# Patient-Reported Shared Decision-Making and Documented Advance Care Planning Among Adults With Alzheimer’s Disease and Related Dementias

**DOI:** 10.64898/2026.10.15.26365674

**Authors:** Xiangren Wang, Tamara G. R. Macieira, Young-Rock Hong, Lee Revere, Zhigang Xie

**Author notes:** Corresponding author: Zhigang Xie, PhD, MBBS, MPA, Assistant Professor, Department of Health Services Research, Management and Policy, College of Public Health and Health Professions, University of Florida PO Box 100195, Gainesville, FL US 32610.

## Abstract

**Objective:** To examine the association between patient-reported shared decision-making (SDM) and documented advance care planning (ACP) among adults with Alzheimer disease and related dementias (ADRD), and to assess whether this association differed between adults with and without ADRD.

**Design:** Retrospective observational study using linked survey and electronic health record (EHR) data.

**Setting and Participants:** Participants aged ≥50 years in the All of Us Research Program who completed a patient-reported SDM measure and had linked EHR records. The primary analytic cohort included 3,635 adults with ADRD; 123,681 adults without ADRD comprised a secondary comparison cohort.

**Methods:** SDM was measured by how often clinicians asked participants about their opinions or preferences regarding medical care or treatment and was categorized as high (always/most of the time) or low (some of the time/ None of the time). Documented ACP was identified within 3 years before or after the SDM assessment. Modified Poisson regression with robust variance estimated prevalence ratios (PRs) and adjusted PRs (aPRs).

**Results:** Among adults with ADRD, 60.6% reported high SDM. Documented ACP prevalence was 5.1% among participants with high SDM and 3.6% among those with low SDM (*P*=0.027). High SDM was associated with greater ACP prevalence in the unadjusted model (PR, 1.44; 95% CI, 1.04-1.99); the association remained positive but was attenuated after full adjustment (aPR, 1.28; 95% CI, 0.93-1.77). Among adults without ADRD, high SDM was associated with documented ACP (aPR, 1.17; 95% CI, 1.03-1.33).

**Conclusions and Implications:** Among adults with ADRD, high SDM was associated with greater ACP prevalence in unadjusted analyses, although the fully adjusted association was not statistically significant. In contrast, higher SDM remained significantly associated with ACP among those without ADRD. The low prevalence of ACP highlights an important opportunity to expand discussions of current and future care preferences as a routine component of patient-centered care.

## Introduction

Advance care planning (ACP) is a structured, ongoing process through which individuals, families, and clinicians discuss, document, and periodically revisit preferences for future medical care in accordance with patients’ values and goals.^1,2^ Beyond supporting patient-centered decision-making, ACP may have important implications for healthcare quality and utilization by improving communication about treatment preferences and preparing patients and care partners for future decisions.^3,4^ This is especially important for people living with Alzheimer disease and related dementias (ADRD), as progressive cognitive impairment can narrow the window during which individuals can directly articulate their values and participate meaningfully in decisions about future care, making timely engagement in preference-sensitive discussions particularly important.^1,3,4^

Prior studies have associated ACP with greater documentation of care preferences, increased hospice use, and, in some populations, lower use of potentially burdensome hospital and intensive care services near the end of life.^5–8^ Reflecting its importance in serious illness care including dementia care, ACP has been incorporated into quality measurement and healthcare delivery initiatives by the Centers for Medicare & Medicaid Services (CMS) and the National Committee for Quality Assurance.^9^ In 2016, CMS began reimbursing clinicians for dedicated ACP discussions through Current Procedural Terminology (CPT) codes 99497 and 99498, creating a standardized mechanism for reimbursing and identifying documented ACP encounters in Medicare.

Despite these efforts, use of documented (billed) ACP services has remained relatively limited. In the first year after Medicare introduced dedicated ACP codes, fewer than 2% of fee-for-service Medicare beneficiaries had a documented ACP encounter; use increased in subsequent years but remained uncommon, with cumulative use reaching approximately 7.5% during 2016 through 2019.^10–13^ Uptake has also varied substantially across clinicians, specialties, care settings, and geographic regions.^14–17^ These patterns likely reflect barriers operating at multiple levels. Clinicians may lack awareness of the new ACP billing codes, have limited time, uncertainty about when or how to initiate ACP, or discomfort discussing future deterioration and end-of-life care.^12,18,19^ Patients and care partners may likewise experience uncertainty, emotional distress, or reluctance to engage in such conversations, particularly when prognosis is uncertain.^20^ Administrative and financial considerations, including documentation requirements and perceptions that reimbursement may not fully compensate for the time and complexity of ACP discussions, may further constrain routine implementation. More broadly, differences in expectations and readiness among patients, care partners, clinicians, and healthcare organizations may make ACP difficult to integrate consistently into usual care.^21,22^

These challenges may be particularly consequential for people living with ADRD due to the nature of the disease.^1,3,4,23^ As care planning in dementia is inherently complex because the course of cognitive and functional decline is variable, treatment decisions often involve family or other care partners, and preferences expressed earlier in the disease trajectory may need to be revisited as health status and care needs evolve.^24–27^ Shared decision-making (SDM) may offer an important pathway for facilitating these conversations. By actively eliciting patients’ values, beliefs, and preferences and incorporating them into treatment decisions, SDM may normalize preference-sensitive discussions during routine care and create opportunities for subsequent ACP before decision-making capacity becomes substantially impaired.^28–30^ A recent systematic review and conceptual model proposed that SDM and ACP may be viewed as complementary components of an iterative decision-making process in which patients, care partners, and clinicians share information, clarify values, and revisit preferences over time.^31^

Although evidence suggests that better SDM is associated with greater ACP engagement, this evidence remains limited in the U.S. context.^32–34^ Trials of decision aids and communication interventions in serious illness suggest that interventions designed to support SDM can improve patient knowledge, preparation for decision-making, and, in some studies, documentation of advance directives or treatment preferences.^32^ A systematic review of SDM tools for noncancer serious illness similarly found that such approaches may improve patient satisfaction with communication and advance directive documentation, although the evidence base was small and heterogeneous.^33^ Much of this literature has evaluated structured interventions rather than naturally occurring patient-clinician interactions, and many studies have focused on cancer, general serious illness, or specific treatment decisions. Consequently, less is known about whether a fundamental element of SDM in routine practice—whether clinicians ask patients about their opinions and preferences regarding medical care—is associated with documented ACP among adults with ADRD.^31^ This distinction is important because preference elicitation is a core, actionable component of patient-centered care and may reflect a broader clinical environment in which discussions about future care preferences are more likely to occur.

Using data from the All of Us Research Program, we examined the association between patient-reported SDM and documented ACP among adults aged 50 years or older with ADRD. We hypothesized that participants who reported that clinicians more consistently elicited their opinions and treatment preferences would have a higher prevalence of documented ACP. As a secondary analysis, we examined the corresponding association among adults without ADRD and formally tested whether the association between SDM and documented ACP differed according to ADRD presence.

## Methods

### Data Source and Study Population

We conducted a retrospective observational study using the All of Us Research Program Controlled Tier Curated Data Repository (CDR), version 8, a national research resource designed to support studies of health and health care in a diverse US population.^35^ The All of Us Research Program integrates multiple sources of participant-level information, including demographic characteristics, participant-completed surveys, electronic health record (EHR) data, physical measurements, and other health-related data. For this study, we linked responses from participant surveys collected during 2018 through 2023 with longitudinal EHR data.^35^ The survey data included measures from the Health Care Access and Utilization survey to assess patient experiences with health care and SDM, while EHR data were used for ADRD diagnosis and ACP procedure codes.

Participants were eligible if they were 50 years or older when completed the survey, had linked EHR data, completed the SDM survey item with a date, and reported a usual source of care on the Health Care Access and Utilization survey. The date of the participant’s first response to the SDM item was defined as the index date (T0). The final analytic sample included 127,316 participants, 3,635 with ADRD for the primary analysis and 123,681 without ADRD for the secondary analysis. ADRD was identified by at least one qualifying ICD-coded EHR diagnosis mapped to standardized concepts prior or on index date following the previous study that used All of Research Program data.^36^

The All of Us Research Program operates under a single central institutional review board, and all participants provided written informed consent for participation, including completion of surveys and linkage of EHR. This secondary analysis used deidentified data accessed through the All of Us Researcher Workbench. The University of Florida Institutional Review Board determined that the study was not human subjects research and was therefore exempt from review.

### Measures

The primary outcome was documented ACP, identified from CPT-coded procedures and Systematized Nomenclature of Medicine Clinical Terms (SNOMED) for ACP, including the first 30 minutes and each additional 30 minutes (CPT codes 99497 and 99498, SNOMED code 713603004),. ACP events were ascertained within a symmetric 3-year window before or after T0. This anchored window replaced an “ever documented” definition used in preliminary analyses, under which most ACP events long preceded the survey (79% occurred before T0, by a median of 2.7 years) across observation periods of up to 17 years (**Supplementary Figure 1**). Because the primary outcome window included ACP events occurring both before and after T0, the resulting estimates were interpreted as contemporaneous associations between reported SDM and documented ACP rather than as prospective or causal effects. Sensitivity analyses used progressively narrower windows of 2 years and 1 year before or after T0.

The primary exposure was patient-reported SDM at baseline, asking how often clinicians asked for the participant’s opinions or beliefs about their medical care or treatment. Responses were dichotomized using a top-2-box approach, with Always or Most of the time categorized as high SDM and Some of the time or None of the time were categorized as low SDM, which served as the reference group.

Covariates were selected a priori based on prior literature and their potential relationships with both SDM and ACP. The fully adjusted model included age at T0, sex at birth, race/ethnicity, educational attainment, household income, marital status, having difficulty dressing or bathing, physical function, and health literacy (using a study-specific binary adaptation of three screening questions: low indicated at least two difficulty flags and high indicated fewer than two; **Table 1**). Any missing item yielded a Missing/Unknown composite. *Statistical Analysis*

**Table 1.** Participant Characteristics by Shared Decision-Making Status Among Adults With Alzheimer Disease and Related Dementias.

|  | Individuals with ADRD n (%) |  |  |
| --- | --- | --- | --- |
| Characteristic | Low SDM<br>1,433 (39.4%) | High SDM<br>2,202 (60.6%) | *P-value*<sup>c</sup> |
| **Age (Years)** |  |  | 0.441 |
| <75 | 980 (68.4) | 1,533 (69.6) |  |
| ≥75 | 453 (31.6) | 669 (30.4) |  |
| **Sex at birth** |  |  | 0.035 |
| Male | 675 (47.1) | 1,117 (50.7) |  |
| Female | 758 (52.9) | 1,085 (49.3) |  |
| **Race/Ethnicity** |  |  | 0.478 |
| Non-Hispanic White | 1,067 (74.5) | 1,621 (73.6) |  |
| Non-Hispanic Black | 114 (8.0) | 182 (8.3) |  |
| Non-Hispanic Other | 73 (5.1) | 113 (5.1) |  |
| Hispanic | 123 (8.6) | 218 (9.9) |  |
| Missing/Unknown | 56 (3.9) | 68 (3.1) |  |
| **Marital Status** |  |  | 0.988 |
| Married/Partnered | 826 (57.6) | 1,265 (57.4) |  |
| Not Married | 596 (41.6) | 920 (41.8) |  |
| Missing/Unknown | 11 (0.8) | 17 (0.8) |  |
| **Education** |  |  | 0.008 |
| < College | 570 (39.8) | 983 (44.6) |  |
| ≥College | 849 (59.2) | 1,190 (54.0) |  |
| Missing/Unknown | 14 (1.0) | 29 (1.3) |  |
| **Annual household income (USD)** |  |  | 0.012 |
| Low (<\$50,000) | 464 (32.4) | 825 (37.5) | |
| Middle (\$50,000-\$99,999) | 379 (26.4) | 511 (23.2) | |
| High (≥\$100,000) | 330 (23.0) | 477 (21.7) | |
| Missing/Unknown | 260 (18.1) | 389 (17.7) |  |
| **Having difficulty dressing or bathing** |  |  | 0.969 |
| Yes | 120 (8.4) | 189 (8.6) |  |
| No | 980 (68.4) | 1,507 (68.4) |  |
| Missing/Unknown | 333 (23.2) | 506 (23.0) |  |
| **Physical Function <sup>a</sup>** |  |  | 0.591 |
| Good | 940 (65.6) | 1,477 (67.1) |  |
| Moderate | 270 (18.8) | 381 (17.3) |  |
| Poor | 212 (14.8) | 331 (15.0) |  |
| Missing/Unknown | 11 (0.8) | 13 (0.6) |  |
| **Health Literacy <sup>b</sup>** |  |  | 0.126 |
| High | 992 (69.2) | 1,454 (66.0) |  |
| Low | 402 (28.1) | 678 (30.8) |  |
| Missing/Unknown | 39 (2.7) | 70 (3.2) |  |

<sup>a</sup> Physical function was assessed using the survey item: “To what extent are you able to carry out your everyday physical activities, such as walking, climbing stairs, carrying groceries, or moving a chair?” Responses were categorized as Good (completely or mostly able), Moderate (moderately able), and Poor (a little able or not at all able). Source item: Overall Health: Everyday Activities.
<sup>b</sup> Health literacy uses the original study-specific binary adaptation of three questions: Health literacy was assessed using a study-specific binary adaptation of three questions from the All of Us Overall Health survey: (1) “How confident are you filling out medical forms by yourself?”; (2) “How often do you have someone help you read health-related materials?”; and (3) “How often do you have problems learning about your medical condition because of difficulty understanding written information?”. One difficulty flag for confidence below Quite a bit, one for any frequency of needing reading help, and one for any frequency of difficulty understanding medical information. Low=at least two flags; High=fewer than two. Any item missing makes the composite Missing/Unknown.
<sup>c</sup> P-values were calculated using Fisher’s exact test. For contingency tables larger than 2×2, P-values were estimated using 100,000 Monte Carlo simulations with a fixed random seed. Missing and unknown categories were included in the distribution tests.
Abbreviations: SDM, shared decision-making

The primary analysis was conducted among participants with ADRD. We first summarized participant characteristics overall and by SDM status (high vs. low) using counts and percentages. Differences in categorical characteristics between SDM groups were evaluated using Fisher exact tests, with Monte Carlo simulation for larger contingency tables as appropriate. To estimate the association between SDM and documented ACP, we used modified Poisson regression with a log link and HC3 robust variance to estimate prevalence ratios (PRs) and adjusted prevalence ratios (aPRs) with 95% CIs. Three sequential models were specified. Model 1 included SDM only. Model 2 additionally adjusted for age, sex at birth, and combined race/ethnicity. Model 3 further adjusted for marital or partnership status, educational attainment, annual household income, activities-of-daily-living impairment, physical function, and health literacy. The same analytic sample was retained across the 3 models within each ADRD group; participants with missing covariate information were retained using missing or unknown categories where applicable.

As a secondary analysis, we repeated the fully adjusted analysis among participants without ADRD and formally evaluated whether the association between SDM and documented ACP differed by ADRD status by including an SDM × ADRD interaction term in a combined model. Sensitivity analyses evaluated the robustness of the primary findings using narrower ACP ascertainment windows of 2 years and 1 year before or after the SDM survey date. All hypothesis tests were 2-sided, and P values <.05 were considered statistically significant. Analyses were conducted within the secure All of Us Researcher Workbench cloud environment.

## Results

### Participant Characteristics

Among 3,635 adults with ADRD, 2,202 (60.6%) reported high SDM and 1,433 (39.4%) reported low SDM. Participants in the high- and low-SDM groups were generally comparable with respect to age, race and ethnicity, marital status, activities-of-daily-living impairment, physical function, and health literacy. However, modest differences were observed by sex, education, and household income. Participants reporting high SDM were slightly more likely to be male (50.7% vs 47.1%), less likely to have completed college or higher education (54.0% vs 59.2%), and more likely to have annual household income below $50,000 (37.5% vs 32.4%) than those reporting low SDM (**Table 1**).

### Shared Decision-Making and Documented Advance Care Planning Among Adults With ADRD

Observed prevalence of documented ACP was 4.5% among adults with ADRD, it was higher among participants with high SDM than among those with low SDM (5.1% vs 3.6%; **Figure 1**). In the unadjusted modified Poisson model, high SDM was associated with a 44% higher prevalence of documented ACP compared with low SDM (PR, 1.44; 95% CI, 1.04-1.99). After adjustment for age, sex, and race and ethnicity, the association was attenuated and the confidence interval included the null (aPR, 1.36; 95% CI, 0.98-1.87). In the fully adjusted model, which additionally accounted for marital status, education, household income, health literacy, having difficulty dressing or bathing, and physical function, the association remained positive but was further attenuated and imprecisely estimated (aPR, 1.28; 95% CI, 0.93-1.77) (**Table 2**).

**Figure 1.**
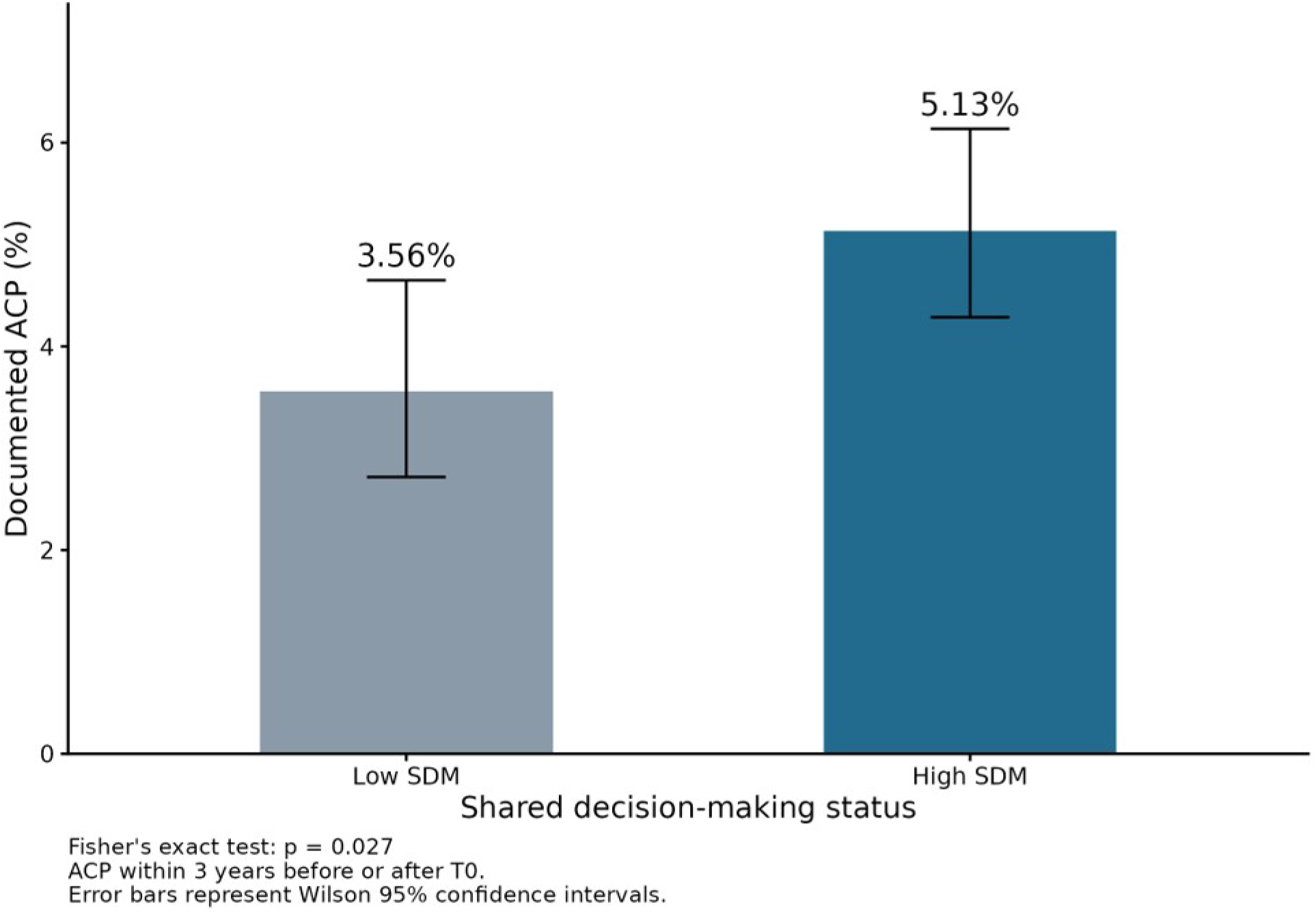
Prevalence of Documented Advance Care Planning by Shared Decision-Making Status Among Adults with Alzheimer Disease and Related Dementias

**Table 2.** Association Between Shared Decision-Making and Documented Advance Care Planning Among Adults With Alzheimer Disease and Related Dementias.

|  | **Individuals with ADRD**<br>**PR/aPR (95% CI); N=3,635** |  |  |
| --- | --- | --- | --- |
| **Characteristic** | **Model 1** | **Model 2** | **Model 3** |
| **SDM** |  |  |  |
| Low | 1.00 (ref) | 1.00 (ref) | 1.00 (ref) |
| High | 1.44 (1.04-1.99) | 1.36 (0.98-1.87) | 1.28 (0.93-1.77) |
| **Age (Years)** | - |  |  |
| <75 | - | 1.00 (ref) | 1.00 (ref) |
| ≥75 | - | 1.47 (1.09-1.99) | 1.54 (1.14-2.10) |
| **Sex at birth** | - |  |  |
| Male | - | 1.00 (ref) | 1.00 (ref) |
| Female | - | 0.17 (0.11-0.26) | 0.16 (0.10-0.24) |
| **Race/Ethnicity** | - |  |  |
| Non-Hispanic White | - | 1.00 (ref) | 1.00 (ref) |
| Non-Hispanic Black | - | 1.39 (0.80-2.42) | 1.08 (0.59-1.96) |
| Non-Hispanic Other | - | 0.94 (0.47-1.88) | 0.79 (0.39-1.59) |
| Hispanic | - | 1.22 (0.73-2.02) | 0.97 (0.58-1.63) |
| Missing/Unknown | - | 0.56 (0.18-1.74) | 0.50 (0.16-1.58) |
| **Marital Status** | - | - |  |
| Married/Partnered | - | - | 1.00 (ref) |
| Not Married | - | - | 1.40 (1.00-1.95) |
| Missing/Unknown | - | - | 1.34 (0.17-10.59) |
| **Education** | - | - |  |
| < College | - | - | 1.00 (ref) |
| ≥College | - | - | 0.80 (0.57-1.13) |
| Missing/Unknown | - | - | NE |
| **Annual household income (USD)** | - | - |  |
| Low (<\$50,000) | - | - | 1.00 (ref) |
| Middle (\$50,000-\$99,999) | - | - | 1.15 (0.78-1.69) |
| High (≥\$100,000) | - | - | 0.59 (0.33-1.03) |
| Missing/Unknown | - | - | 0.58 (0.35-0.95) |
| **Having difficulty dressing or bathing** | - | - |  |
| Yes | - | - | 1.00 (ref) |
| No | - | - | 0.87 (0.56-1.34) |
| Missing/Unknown | - | - | 0.53 (0.31-0.93) |
| **Physical Function** | - | - |  |
| Good | - | - | 1.00 (ref) |
| Moderate | - | - | 1.73 (1.18-2.54) |
| Poor | - | - | 2.17 (1.46-3.24) |
| Missing/Unknown | - | - | 3.39 (1.05-10.98) |
| **Health Literacy** | - | - |  |
| High | - | - | 1.00 (ref) |
| Low | - | - | 1.15 (0.83-1.60) |
| Missing/Unknown | - | - | 1.25 (0.57-2.77) |

Several covariates were also associated with documented ACP in the fully adjusted ADRD model. Adults aged 75 years or older had a higher prevalence of documented ACP than those younger than 75 years (aPR, 1.54; 95% CI, 1.14-2.10), whereas women had a substantially lower prevalence than men (aPR, 0.16; 95% CI, 0.10-0.24). Moderate and poor physical function were associated with higher ACP prevalence compared with good physical function (moderate: aPR, 1.73; 95% CI, 1.18-2.54; poor: aPR, 2.17; 95% CI, 1.46-3.24). No clear associations were observed for race and ethnicity, education, or health literacy in the fully adjusted model.

### Comparison With Adults Without ADRD

Among 123,681 adults without ADRD (**Supplementary Table 1**), high SDM was also associated with a higher prevalence of documented ACP. In the fully adjusted model, participants reporting high SDM had a 17% higher prevalence of documented ACP than those reporting low SDM (aPR, 1.17; 95% CI, 1.03-1.33) (**Table 3**). The corresponding observed ACP prevalence was 0.9% among participants with high SDM and 0.8% among those with low SDM (**Supplementary Figure 2**). The multiplicative interaction between SDM and ADRD status was not statistically significant (*P*=.523).

**Table 3.** Association Between Shared Decision-Making and Documented Advance Care Planning Among Adults Without Alzheimer Disease and Related Dementias.

|  | **Individuals without ADRD**<br>**PR/aPR (95% CI); N=123,681** |  |  |
| --- | --- | --- | --- |
| **Characteristic** | **Model 1** | **Model 2** | **Model 3** |
| **SDM** |  |  |  |
| Low | 1.00 (ref) | 1.00 (ref) | 1.00 (ref) |
| High | 1.23 (1.09-1.40) | 1.15 (1.02-1.31) | 1.17 (1.03-1.33) |
| **Age (Years)** | - |  |  |
| <75 | - | 1.00 (ref) | 1.00 (ref) |
| ≥75 | - | 1.82 (1.59-2.08) | 1.85 (1.61-2.12) |
| **Sex at birth** | - |  |  |
| Male | - | 1.00 (ref) | 1.00 (ref) |
| Female | - | 0.23 (0.20-0.26) | 0.19 (0.17-0.22) |
| **Race/Ethnicity** | - |  |  |
| Non-Hispanic White | - | 1.00 (ref) | 1.00 (ref) |
| Non-Hispanic Black | - | 1.29 (1.05-1.59) | 0.88 (0.71-1.10) |
| Non-Hispanic Other | - | 1.18 (0.91-1.51) | 1.06 (0.82-1.37) |
| Hispanic | - | 1.23 (1.00-1.52) | 0.93 (0.74-1.16) |
| Missing/Unknown | - | 1.69 (1.26-2.26) | 1.45 (1.08-1.95) |
| **Marital Status** | - | - |  |
| Married/Partnered | - | - | 1.00 (ref) |
| Not Married | - | - | 1.41 (1.23-1.61) |
| Missing/Unknown | - | - | 1.36 (0.83-2.23) |
| **Education** | - | - |  |
| < College | - | - | 1.00 (ref) |
| ≥College | - | - | 0.79 (0.69-0.91) |
| Missing/Unknown | - | - | 0.69 (0.38-1.26) |
| **Annual household income (USD)** | - | - |  |
| Low (<\$50,000) | - | - | 1.00 (ref) |
| Middle (\$50,000-\$99,999) | - | - | 0.86 (0.73-1.01) |
| High (≥\$100,000) | - | - | 0.44 (0.36-0.55) |
| Missing/Unknown | - | - | 0.61 (0.49-0.75) |
| **Having difficulty dressing or bathing** | - | - |  |
| Yes | - | - | 1.00 (ref) |
| No | - | - | 0.67 (0.53-0.85) |
| Missing/Unknown | - | - | 0.34 (0.25-0.45) |
| **Physical Function** | - | - |  |
| Good | - | - | 1.00 (ref) |
| Moderate | - | - | 1.69 (1.42-2.00) |
| Poor | - | - | 1.75 (1.41-2.17) |
| Missing/Unknown | - | - | 1.72 (0.94-3.14) |
| **Health Literacy** | - | - |  |
| High | - | - | 1.00 (ref) |
| Low | - | - | 0.97 (0.83-1.14) |
| Missing/Unknown | - | - | 1.10 (0.77-1.59) |

### Sensitivity Analyses

Results were generally similar when narrower ACP ascertainment windows were used, although effect estimates moved closer to the null and confidence intervals widened. Among adults with ADRD, the fully adjusted aPR for high vs low SDM was 1.27 (95% CI, 0.89-1.82) using a ±2-year window, and 1.11 (95% CI, 0.71-1.75) using a ±1-year window. Among adults without ADRD, corresponding estimates were 1.13 (95% CI, 0.98-1.30), and 1.10 (95% CI, 0.93-1.31), respectively (**Supplementary Table 2**).

## Discussion

In this national analysis of adults with ADRD, patient-reported SDM was positively associated with documented ACP, although the fully adjusted estimate became statistically nonsignificant. Participants reporting high SDM had a higher observed prevalence of documented ACP than those reporting low SDM, and the association remained in the same direction after adjustment for demographic, socioeconomic, functional, and health-literacy characteristics. A similar, somewhat smaller association was observed among adults without ADRD, and the SDM × ADRD interaction was not statistically significant, providing no evidence that the relative association differed by ADRD status. Together, these findings suggest that clinician elicitation of patients’ opinions and treatment preferences and formal ACP documentation may represent related components of a broader patient-centered care environment, rather than a relationship unique to dementia.

Our findings are consistent with conceptual models that position SDM and ACP as related components of an iterative patient-centered decision-making process.^31^ SDM primarily supports decisions about current care by eliciting patients’ values, concerns, and preferences, whereas ACP extends those discussions to future circumstances in which decision-making capacity may be diminished. Rosca and colleagues proposed that these processes may be integrated over time, with preference elicitation during routine treatment decisions providing a foundation for broader discussions of future goals of care.^31^ In our study, because ACP events were assessed within a symmetric window surrounding the SDM survey, these findings should not be interpreted as evidence that SDM caused subsequent ACP. Rather, they suggest that preference elicitation and formal ACP documentation may cluster within a broader patient-centered care environment.

The low prevalence of documented ACP in our ADRD cohort is consistent with prior evidence that formal ACP engagement remains limited in dementia care, even when ACP is defined more broadly than billing records alone.^10–13^ Studies of older adults with dementia have shown that many patients do not complete or document ACP before decisional capacity declines, and ACP discussions are often initiated relatively late in the disease trajectory.^37–40^ This pattern is clinically important because opportunities for direct participation may narrow as cognitive and functional impairment progresses, increasing reliance on surrogate decision-makers and retrospective interpretation of patients’ preferences. The relatively low prevalence observed in our study therefore likely reflects not only the limited diffusion of dedicated ACP billing but also the broader difficulty of integrating future-care discussions into routine dementia care. At the same time, our estimate should be interpreted specifically as the prevalence of documented ACP, not the prevalence of all ACP-related activity. Patients may discuss future care preferences, complete advance directives, or engage in family- or clinician (including nurses and other allied health professions)-led planning without formal documentation.^41–45^ Thus, the gap between the clinical importance of ACP and its documentation in routine care may be even more nuanced than billing data alone can capture.

The positive association between SDM and documented ACP was also present among adults without ADRD, and we found no evidence that ADRD status modified this association. This pattern is consistent with literature suggesting that preference elicitation and ACP are complementary components of patient-centered decision-making across serious illnesses rather than processes specific to dementia.^33^ Nevertheless, the clinical importance of SDM may be heightened in ADRD because progressive cognitive decline can narrow the period during which patients can directly express their values and participate in future care decisions.^23^ Dementia-specific reviews and recommendations therefore emphasize early, iterative engagement and continued involvement of patients and care partners as decision-making capacity changes over time.^39,40^

The attenuation of the SDM–ACP association after covariate adjustment suggests that part of the observed relationship may reflect broader differences in patient characteristics and care context rather than SDM alone. In the ADRD cohort, the prevalence ratio decreased from 1.44 in the unadjusted model to 1.28 after adjustment for demographic, socioeconomic, functional, and health-literacy factors, while remaining in the same positive direction. This pattern suggests that SDM and ACP may be embedded within a wider set of patient and care-process characteristics that influence both preference elicitation and ACP documentation.^31^

Several limitations should also be considered. First, ACP was identified within a symmetric window before and after the SDM survey, precluding temporal or causal inference. Second, SDM was measured with a single patient-reported item and therefore does not capture all dimensions of SDM. Third, CPT-based ACP ascertainment likely underestimates broader ACP activity because conversations, advance directive completion, and care-planning discussions may occur without generating dedicated ACP billing codes. Fourth, ADRD was identified from EHR diagnoses and may be subject to misclassification, and residual confounding from unmeasured clinical or care-system factors remains possible. Finally, participation in All of Us and completion of the relevant surveys may limit generalizability to the broader US population with ADRD.

## Conclusions

In this national analysis, greater patient-reported SDM was associated with a higher prevalence of documented ACP among adults with ADRD, although the fully adjusted estimate was statistically nonsignificant. Similar findings among adults without ADRD and the absence of a significant interaction suggest that preference elicitation and ACP may reflect related components of patient-centered care across populations. Given the progressive loss of decision-making capacity in dementia, integrating routine elicitation of patient preferences with earlier and ongoing ACP may help support more person-centered care throughout the disease trajectory.

## Disclosures and Acknowledgments

### Author Contributions

X.W.: conceptualization; methodology; software; formal analysis; original draft; writing— review and editing. T.M.: conceptualization; writing—review and editing. Y.H.: conceptualization; writing—review and editing. L.R.: conceptualization; writing—review and editing. Z.X.: conceptualization; methodology; original draft; supervision; writing—review and editing; corresponding author.

### Ethical Considerations

As the All of Us Research Program Data are publicly available through an institutional data use agreement and provided as a de-identified dataset, the University of Florida Institutional Review Board determined that this study was nonhuman subject research.

## Data Availability

Data are available to approved researchers through the All of Us Researcher Workbench (Controlled Tier); the analytic code is available from the corresponding author upon reasonable request.

https://www.researchallofus.org/

## Acknowledgments

The All of Us Research Program is supported by the National Institutes of Health, Office of the Director. We gratefully acknowledge All of Us participants for their contributions, without whom this research would not have been possible, and the National Institutes of Health’s All of Us Research Program for making available the participant data examined in this study.

