## Supplementary Material for "Patient-Reported Shared Decision-Making and Documented Advance Care Planning Among Adults With Alzheimer’s Disease and Related Dementias"

Contents: Supplementary Table 1; Supplementary Table 2; Supplementary Figure 1; Supplementary Figure 2

**Supplementary Table 1.** Participant Characteristics by shared decision making among individuals without ADRD

| | Individuals without ADRD n (%) | | |
|---|---|---|---|
| **Characteristic** | **Low SDM 46,870 (37.9%)** | **High SDM 76,811 (62.1%)** | **P** |
| **Age (Years)** | | | **0.025** |
| <75 | 40,601 (86.6) | 66,191 (86.2) | |
| ≥75 | 6,269 (13.4) | 10,620 (13.8) | |
| **Sex at birth** | | | **<0.001** |
| Male | 16,437 (35.1) | 30,068 (39.1) | |
| Female | 30,433 (64.9) | 46,743 (60.9) | |
| **Race/Ethnicity** | | | **<0.001** |
| Non-Hispanic White | 34,913 (74.5) | 55,742 (72.6) | |
| Non-Hispanic Black | 4,084 (8.7) | 7,543 (9.8) | |
| Non-Hispanic Other | 2,674 (5.7) | 4,314 (5.6) | |
| Hispanic | 3,964 (8.5) | 7,340 (9.6) | |
| Missing/Unknown | 1,235 (2.6) | 1,872 (2.4) | |
| **Marital Status** | | | **<0.001** |
| Married/Partnered | 28,885 (61.6) | 48,681 (63.4) | |
| Not Married | 17,449 (37.2) | 27,331 (35.6) | |
| Missing/Unknown | 536 (1.1) | 799 (1.0) | |
| **Education** | | | **<0.001** |
| < College | 18,171 (38.8) | 30,984 (40.3) | |
| ≥College | 28,197 (60.2) | 44,889 (58.4) | |
| Missing/Unknown | 502 (1.1) | 938 (1.2) | |

| | | | |
|---|---|---|---|
| **Annual household income (USD)** | | | **0.762** |
| Low (<$50,000) | 12,994 (27.7) | 21,306 (27.7) | |
| Middle ($50,000-$99,999) | 11,998 (25.6) | 19,700 (25.6) | |
| High (≥$100,000) | 15,903 (33.9) | 25,882 (33.7) | |
| Missing/Unknown | 5,975 (12.7) | 9,923 (12.9) | |
| **Having difficulty dressing or bathing** | | | **<0.001** |
| Yes | 1,701 (3.6) | 2,355 (3.1) | |
| No | 36,977 (78.9) | 61,225 (79.7) | |
| Missing/Unknown | 8,192 (17.5) | 13,231 (17.2) | |
| **Physical Function** | | | **<0.001** |
| Good | 37,439 (79.9) | 62,458 (81.3) | |
| Moderate | 5,546 (11.8) | 8,583 (11.2) | |
| Poor | 3,617 (7.7) | 5,291 (6.9) | |
| Missing/Unknown | 268 (0.6) | 479 (0.6) | |
| **Health Literacy** | | | **0.019** |
| High | 38,465 (82.1) | 62,802 (81.8) | |
| Low | 7,472 (15.9) | 12,300 (16.0) | |
| Missing/Unknown | 933 (2.0) | 1,709 (2.2) | |

**Supplementary Table 2.** Association Between Shared Decision-Making and Documented Advance Care Planning Using Alternative Ascertainment Windows

| Advance care Planning ascertainment window (Full Model) | **Individuals with ADRD aPR (95% CI)** | **Individuals without ADRD aPR (95% CI)** |
|---|---|---|
| +/-2 years | 1.27 (0.89-1.82) | 1.13 (0.98-1.30) |
| +/-1 year | 1.11 (0.71-1.75) | 1.10 (0.93-1.31) |

**Supplementary Figure 1.** Distribution of Documented Advance Care Planning by Shared Decision-Making Status Among Adults with and Without Alzheimer Disease and Related Dementias (ADRD)

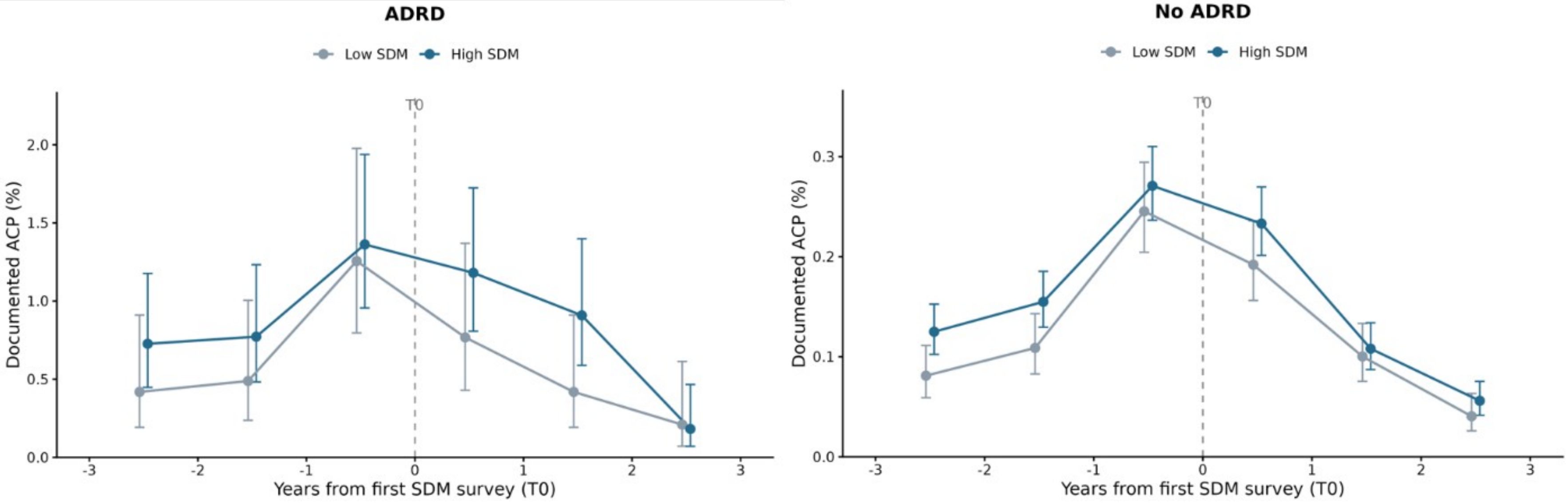

**Supplementary Figure 2.** Prevalence of Documented Advance Care Planning by Shared Decision-Making Status Among Adults Without Alzheimer Disease and Related Dementias

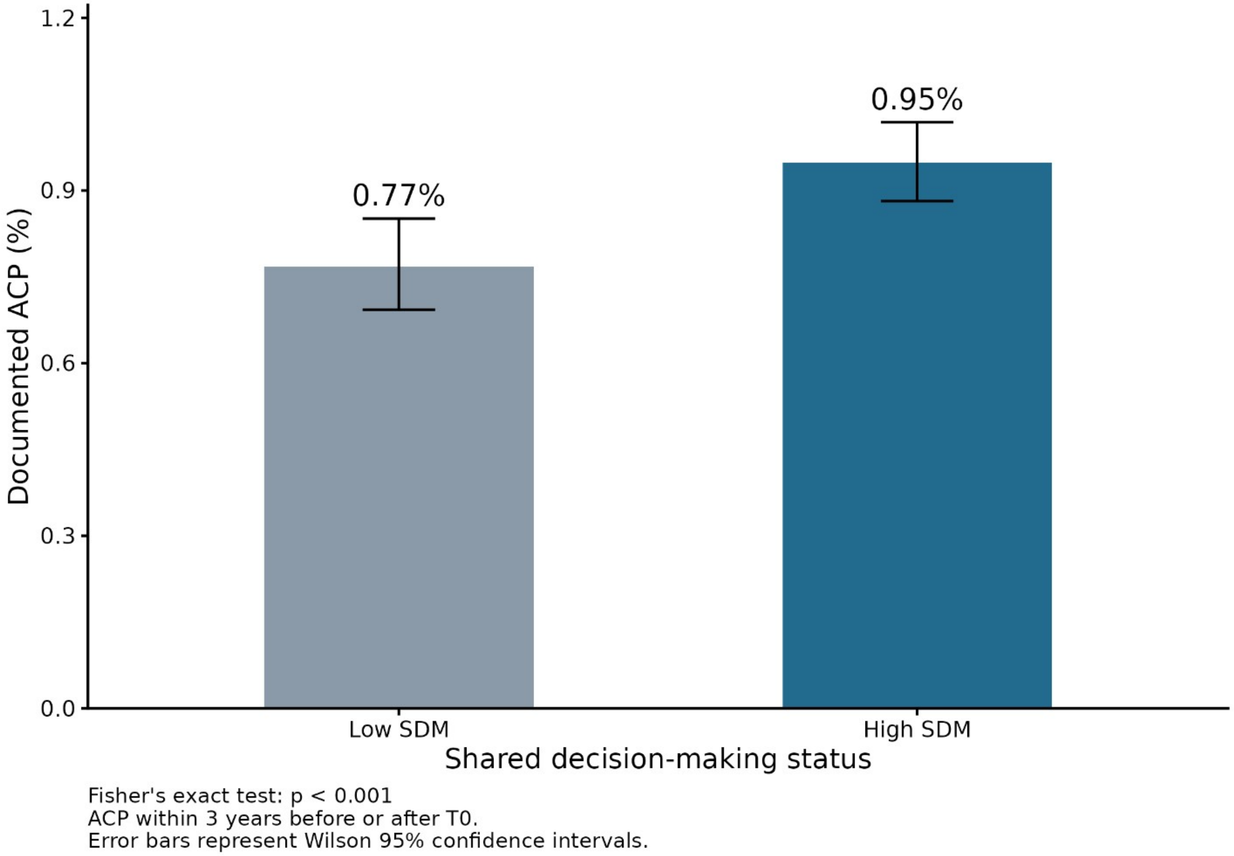
